# Cue-Induced Retrieval and Reconsolidation with Episodic Future Thinking for Craving and Delay Discounting in Opioid Use Disorder: A Pilot Randomized Controlled Trial

**DOI:** 10.64898/2026.08.19.26360819

**Authors:** Matin Toulami, Keyvan Ghasemi, Parnian Rafei, Jasmin Vassileva, Mohammad Salehi, Hamed Ekhtiari, Tara Rezapour

## Abstract

**Aims:** To evaluate whether Cue-Induced Retrieval and Reconsolidation with Episodic Future Thinking (CIREF), which combines personalized drug-cue retrieval with structured future-oriented processing, produces greater changes in craving and delay discounting than a recent-past episodic active control in individuals with opioid use disorder receiving methadone maintenance treatment.

**Design:** Multicentre, two-arm, parallel-group randomized controlled pilot trial with per-protocol analyses.

**Setting:** Two outpatient addiction treatment and rehabilitation centres in Tehran, Iran.

**Participants:** Thirty participants with opioid use disorder receiving methadone maintenance treatment were randomized to CIREF or Episodic Recent Thinking (ERT; n = 15 per group). Twenty-eight completed the intervention and were included in the analyses (n = 14 per group).

**Intervention and comparator:** Participants completed one screening, baseline, and personalized cue-development session followed by three 75-minute intervention sessions. CIREF combined personalized drug-cue retrieval with future-oriented simulation, prediction, intention, and planning. ERT was structurally matched but anchored episodic processing to the recent past.

**Measurements:** Primary craving outcomes comprised the three Desire for Drug Questionnaire (DDQ) subscales assessing session-level phasic/current craving immediately before and after each intervention session and the four Obsessive–Compulsive Drug Use Scale (OCDUS) subscales assessing tonic craving before and after the intervention period. The secondary outcome was Monetary Choice Questionnaire (MCQ) log(k), with more negative values indicating less steep delay discounting.

**Findings:** After Holm correction across the three DDQ subscales, Group × Occasion interactions indicated greater reductions with CIREF for Desire and Intention to Drug Use, FGG(1.23, 32.09) = 24.37, pHolm < .001, η²p = .484, and Negative Reinforcement, FGG(1.35, 35.23) = 17.67, pHolm < .001, η²p = .405, but not Drug Abuse Control (pHolm = .172). After Holm correction across the four OCDUS subscales, only Desire and Mental Preoccupation with Drugs showed a significant Group × Time interaction, F(1, 26) = 12.35, pHolm = .007, η²p = .322; the remaining subscales were not significant (adjusted ps ≥ .177). MCQ log(k) showed a Group × Time interaction, F(1, 26) = 7.18, p = .013, η²p = .217; mean log(k) changed from −1.22 (0.36) to −1.80 (0.55) in CIREF and from −1.39 (0.32) to −1.44 (0.44) in ERT.

**Conclusions:** In this small pilot sample, the future-oriented retrieval-based intervention produced greater changes than the recent-past active control in two dimensions of session-level phasic craving, one dimension of tonic craving, and monetary delay discounting. The results are preliminary and do not establish memory reconsolidation or effects on relapse or longer-term clinical outcomes.

## INTRODUCTION

*“What matters in life is not what happens to you but what you remember and how you remember it”*

— Gabriel García Márquez, Living to Tell the Tale (2002)

Opioid use disorder (OUD) is a chronic, relapsing condition characterized by persistent opioid use and a sustained vulnerability to relapse (1). Repeated drug use establishes persistent drug-related memories that can be reactivated by drug-associated cues and contribute to craving and relapse (2,3). Methadone maintenance treatment (MMT) is among the most effective treatments for OUD and is associated with reduced illicit opioid use, mortality, and other opioid-related harms (1,4). Nevertheless, drug-related cues and craving remain clinically relevant during MMT, as salient internal or environmental cues may retrieve established drug-related memories and promote renewed drug-seeking (3,5–8). This vulnerability is important because the protective benefits of MMT are greatest during sustained treatment, whereas discontinuation can increase the risk of mortality and other adverse outcomes (1,4). Adjunctive behavioural interventions targeting persistent drug-cue memories and related cognitive and motivational processes may therefore complement MMT and support long-term recovery (5,9,10).

Drug-related cues associated with previous opioid use can acquire heightened motivational salience, capture attention, retrieve drug-related memories, and elicit conditioned emotional and behavioural responses (6,10,11). Such cue reactivity can intensify craving and promote drug-seeking behaviour, including among individuals receiving MMT and after prolonged periods of abstinence (5–8). Cue-exposure therapy (CET) has been used to reduce conditioned responses by repeatedly presenting drug-related cues in the absence of drug use or reinforcement, thereby promoting extinction learning (12,13). However, clinical effects have been inconsistent, and reductions achieved during treatment may not generalize reliably to drug cues encountered in everyday contexts (12,13). Extinction is thought to establish new inhibitory learning that competes with, rather than eliminates, the previously acquired cue–drug association, leaving conditioned responses vulnerable to re-emergence outside the extinction context (13,14). These limitations have motivated approaches aimed at modifying reactivated drug-related memories following retrieval (2,13–15).

Memory reconsolidation provides a framework for such retrieval-dependent modification (2,15). When a consolidated memory is reactivated, it may enter a transiently labile state during which new information can be incorporated before the memory restabilizes (16,17). However, retrieval does not necessarily induce memory destabilization; whether reconsolidation is engaged depends on boundary conditions related to both the memory and the retrieval experience (18,19). Prediction error reflects a mismatch between expected and actual outcomes during retrieval and has been proposed as an important trigger for destabilization, although its necessity and optimal magnitude remain uncertain (20–22). Reconsolidation should therefore be viewed as a conditional rather than automatic consequence of retrieval (18,21). When the relevant conditions are met, the post-retrieval period may provide an opportunity to introduce new information capable of altering the affective or motivational properties of the reactivated memory (2,15,19). A central question is consequently what form of information might provide clinically useful updating content following retrieval.

Episodic future thinking (EFT) represents a potential source of such adaptive information (13). EFT involves the simulation of personally relevant events that may occur in one’s future (23). By increasing the salience and psychological accessibility of future alternatives, EFT may enhance the subjective value of delayed outcomes and reduce delay discounting, thereby shifting intertemporal choice away from smaller immediate rewards toward longer-term outcomes (23–25). Experimental studies have repeatedly demonstrated reductions in delay discounting with EFT, in individuals with alcohol, tobacco, and cocaine use, whereas evidence for effects on craving, drug demand, and actual substance use is more limited and heterogeneous (26–30). Within a retrieval-based framework, future simulations may additionally provide novel, personally meaningful prospective information following drug-memory retrieval that could become associated with reactivated drug-related representations and potentially alter their motivational significance (13).

Future-oriented cognition may be particularly relevant to OUD. Studies of individuals with chronic opioid use have shown impairments in episodic foresight (31,32), while shortened time horizons and reduced sensitivity to future consequences have also been reported in individuals with heroin dependence (33). Episodic foresight and delay discounting are conceptually distinct processes, but each may reduce the influence of delayed or anticipated future outcomes on present decision-making (23,34). This may be especially relevant during MMT, for which sustained engagement is associated with better clinical outcomes (1,4). By making personally meaningful future outcomes more concrete and accessible, EFT may increase their influence on current choices and broaden the temporal window over which consequences are considered (23,25,29,35). However, direct evidence evaluating EFT specifically in individuals with OUD receiving MMT remains sparse (26).

Building on these line of research, Rafei et al. (13) proposed the CIREF conceptual framework, integrating personalized cue-induced retrieval with structured future-oriented cognition within a reconsolidation-informed model. In the present study, this framework was operationalized as Cue-Induced Retrieval and Reconsolidation with Episodic Future Thinking (CIREF). Exposure to personally relevant drug cues is intended to retrieve associated drug-related memories, which may become susceptible to modification if appropriate reconsolidation boundary conditions are met (13,18,19). Participants subsequently construct future situations involving these cues and consider alternative responses and outcomes. Within this framework, EFT provides personally relevant, future-oriented information that can be introduced following retrieval and may potentially become associated with reactivated drug-related representations (13). Importantly, EFT is conceptualized as the content for potential updating rather than as the mechanism that produces prediction error, memory destabilization, or reconsolidation itself (13,20–22).

The CIREF framework draws on Szpunar et al.’s taxonomy of future-oriented cognition, which distinguishes four related processes: simulation, prediction, intention, and planning (36). Simulation involves constructing a specific future event; prediction involves anticipating possible responses and consequences; intention involves forming a desired future goal; and planning involves organizing small, concrete steps toward goal attainment (36). Together, these processes may support self-projection, enhance the accessibility of future outcomes, and facilitate goal-directed responses to drug-related cues (13,36,37). Although retrieval-based memory interventions and EFT have each been studied independently, their integration within a structured intervention combining personalized drug-cue retrieval with future-oriented episodic processing has not, to our knowledge (13), been empirically evaluated. It therefore remains unknown whether CIREF produces different effects on craving and delay discounting than pairing the same retrieval procedure with recent-past episodic thinking.

The present multicentre pilot randomized controlled trial provides the first empirical evaluation of CIREF in adults with OUD receiving MMT. CIREF was compared with Episodic Recent Thinking (ERT), an active control condition matched on personalized cue exposure, therapist contact, episodic elaboration, session duration, and the general sequence of intervention exercises, but anchored to recent-past rather than future-oriented episodic processing. The primary outcomes comprised dimensions of phasic craving, assessed before and after each intervention session, and tonic craving, assessed before and after the intervention period; delay discounting was the secondary outcome.

## METHODS

### Study Design

This pilot study was a multicentre, two-arm, parallel-group randomized controlled trial designed to evaluate the preliminary efficacy of the CIREF intervention in individuals with opioid use disorder (OUD) receiving methadone maintenance treatment (MMT). Participants were allocated to either CIREF or the active control condition, Episodic Recent Thinking (ERT). The study protocol was developed in accordance with the SPIRIT guidelines (38). The trial was registered in the Iranian Registry of Clinical Trials (IRCTID: IRCT20210804052079N1) and registered through the Open Science Framework (OSF) registries (OSF Preregistration; DOI: 10.17605/OSF.IO/GQTP7). The IRCT registration was subsequently updated to reflect amendments to the protocol and study procedures. Study documentation, historical protocol versions, protocol amendments and deviations, and supporting materials are publicly available through the Open Science Framework project repository (DOI: 10.17605/OSF.IO/Q6W9D). No interim analyses or formal stopping rules were planned, and no data monitoring committee (DMC) was established because the trial was considered minimal risk.

The study comprised four weekly, one-on-one, in-person sessions: one screening, baseline-assessment, and personalized cue-development session, followed by three 75- minute intervention sessions. Each intervention session was delivered according to a standardized manualized protocol. Intervention fidelity was monitored by TR and KGH through session recordings, attendance and timing documentation, session notes, and structured supervision to support adherence to the protocol and identify procedural deviations; adherence was not quantitatively scored. Supervision also supported consistent implementation of the condition-specific procedures, namely future-oriented episodic processing in CIREF and recent-past episodic processing in ERT. All study procedures were conducted in private rooms at the participating clinics to minimize environmental variability.

### Sample Size

Because this was a pilot trial, the study was not powered to provide a definitive test of treatment efficacy. The prespecified randomized sample target was 30 participants, with 15 allocated to each condition, to provide preliminary estimates of intervention effects and inform the design of a subsequent adequately powered trial. The achieved randomized sample was 30 participants.

### Participants

Participants were recruited from two outpatient addiction treatment and rehabilitation centres in Tehran, Iran, providing pharmacological and psychological care for individuals with SUD under an identical MMT protocol (39). Recruitment was conducted sequentially across the two centres, with identical recruitment, screening, randomization, intervention, and assessment procedures used at both sites. Sixteen participants were randomized at Centre 1 and 14 at Centre 2. Recruitment was facilitated by clinic staff distributing study information leaflets, followed by a private explanation of the study procedures by the research team to support informed and voluntary participation.

The inclusion criteria were: (1) age 18–50 years; (2) a DSM-5 diagnosis of OUD (40), documented in the clinic medical/IDATIS record following clinical assessment at MMT initiation; (3) receipt of MMT for at least three months at a daily methadone dose of 60– 180 mg; (4) at least eight years of formal education to ensure adequate comprehension of the intervention materials; and (5) ability and willingness to provide written informed consent.

Eligibility screening additionally required an education-adjusted Montreal Cognitive Assessment (MoCA) score ≥25, with one additional point applied for participants with fewer than 12 years of education, a Beck Depression Inventory-II (BDI-II) score <14 (41), and a Snaith-Hamilton Pleasure Scale (SHAPS) score <3 (42). Clinically relevant anhedonia was excluded because it could interfere with participants’ ability to generate positive and concrete intentions during the intervention (28). Exclusion criteria were: (1) a history of neurological disorders (e.g., epilepsy); (2) moderate-to-severe traumatic brain injury with loss of consciousness exceeding 30 minutes; (3) major psychiatric disorders that could interfere with study procedures (e.g., active psychotic disorder); and (4) a current active substance use disorder other than OUD, except nicotine use disorder. Non-opioid substance use was screened using a urine drug dipstick test.

Ability to generate personalized drug-related cues and completion of the core intervention components were evaluated after randomization as aspects of intervention engagement and completion and were not used as pre-randomization eligibility criteria. Of 53 individuals assessed for eligibility, 30 met the eligibility criteria, provided written informed consent, and were randomized (Figure 1).

**Figure 1.**
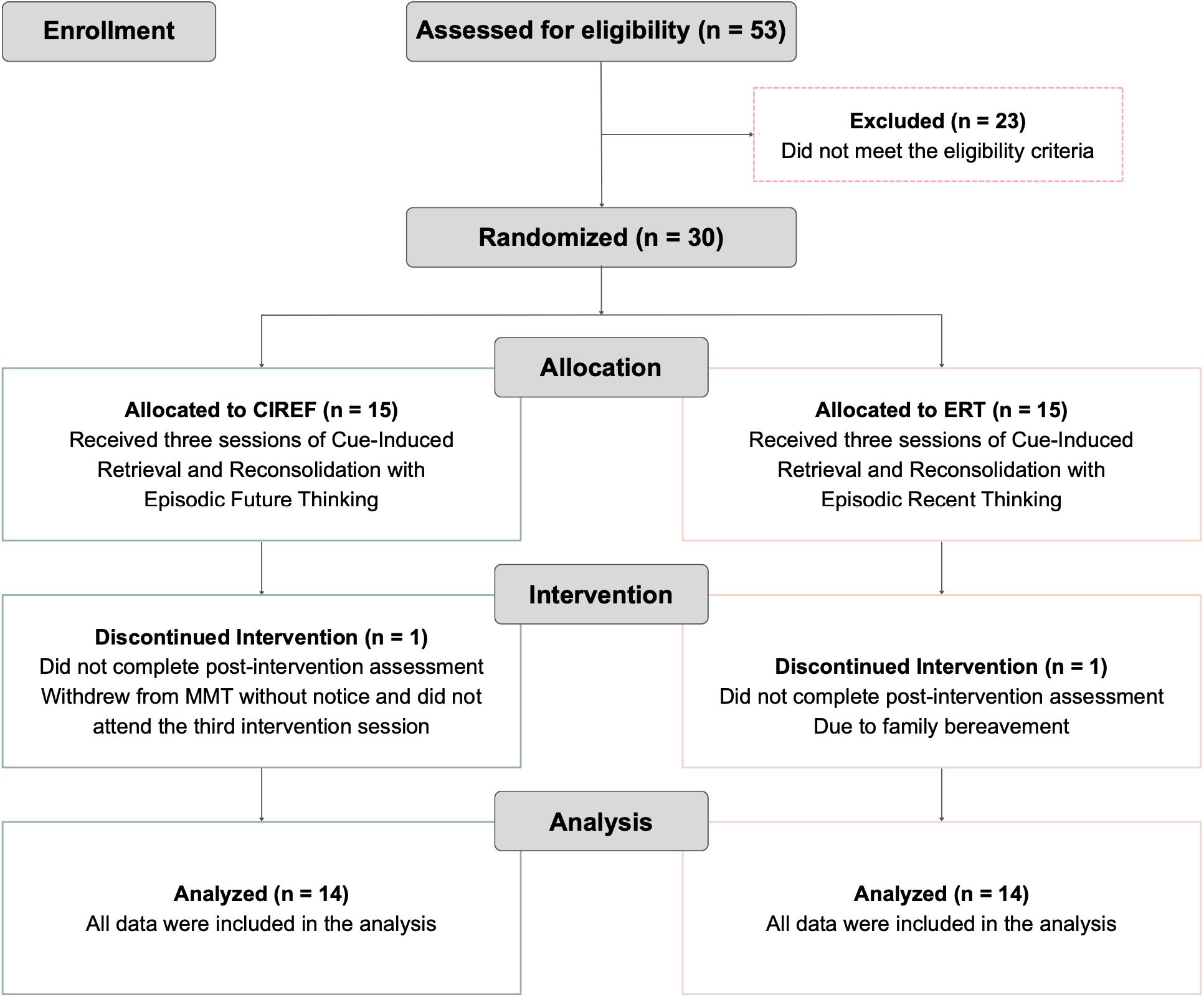
CONSORT flow diagram of participant recruitment, allocation, intervention completion, and analysis. Fifty-three individuals were assessed for eligibility, of whom 23 were excluded for not meeting the eligibility criteria. Thirty participants were randomized to Cue-Induced Retrieval and Reconsolidation with Episodic Future Thinking (CIREF; n = 15) or Episodic Recent Thinking (ERT; n = 15). One participant in each group discontinued the intervention before completing the post-intervention assessment. The CIREF participant withdrew from methadone maintenance treatment without notice and did not attend the third intervention session; the ERT participant discontinued because of family bereavement. Twenty-eight participants were included in the per-protocol analyses (CIREF, n = 14; ERT, n = 14). **CIREF** = Cue-Induced Retrieval and Reconsolidation with Episodic Future Thinking; **ERT** = Episodic Recent Thinking; MMT = methadone maintenance treatment.

Participants were informed that their decision to participate or withdraw, and the information they provided, would have no bearing on the treatment, or care they received at the clinic and that they could withdraw at any time without penalty. Participants received a gift card for completing study assessments and intervention sessions, redeemable for clinical services provided by the participating centres, including take-home methadone or a 30-minute consultation with a psychologist. The study was approved by the Ethics Committees of the University of Tehran and the Institute for Cognitive Science Studies (Approval Code: IR.UT.IRICSS.REC.1400.011).

### Randomization and Allocation

After completion of eligibility assessment, all 30 eligible participants provided written informed consent and were randomized to CIREF or ERT before the intervention phase began. Randomization was implemented separately within each study site using computer-generated permuted blocks with a fixed block size of four and a 1:1 allocation ratio. Separate allocation sequences were generated for participants recruited at each centre. For each complete block, one of the six possible balanced permutations containing two CIREF and two ERT assignments was used to determine allocation (43). At centre 2, where 14 participants were randomized, the final block was incomplete because recruitment ended before completion of the fourth block. The randomization sequences were generated in Python (version 3.10) by a statistician who was not involved in participant recruitment, enrolment, or intervention delivery. Allocation concealment was maintained using sequentially numbered assignment codes.

Thirty participants were randomized in total (Centre 1, n = 16; Centre 2, n = 14), resulting in equal overall allocation to CIREF (n = 15) and ERT (n = 15). One participant in each group discontinued the intervention before completing the post-intervention assessment, resulting in 28 completers (CIREF, n = 14; ERT, n = 14). The primary analyses were conducted on a per-protocol basis in these 28 participants, with all available data from the included participants retained in the analyses.

### Blinding

Because the intervention required participants to engage in either future-oriented episodic processing in CIREF or recent-past episodic processing in ERT, participants could not be blinded to the temporal orientation of the tasks they performed. However, they were not informed which condition was designated as the experimental intervention or active control and were not informed of the study hypotheses. Intervention providers were aware of allocation because they delivered condition-specific procedures, whereas routine clinic staff remained blinded to group allocation.

### Screening

#### Demographics and clinical characteristics

Demographic characteristics, including age, education, and occupational status, and clinical characteristics, including opioid-use history, route of administration, primary opioid used, and methadone dose, were collected using a structured interview.

#### Eligibility screening

Depressive symptoms, anhedonia, and cognitive functioning were assessed for eligibility using the Beck Depression Inventory-II (BDI-II) (41,44), Snaith–Hamilton Pleasure Scale (SHAPS) (42,45), and Montreal Cognitive Assessment (MoCA) (46–48), respectively. Eligibility required a BDI-II score <14, a SHAPS score <3, and an education-adjusted MoCA score ≥25, with one additional point applied for participants with fewer than 12 years of education. After eligibility had been established, the SHAPS and MoCA were administered again as baseline measures only; these baseline reassessments did not determine or reassess eligibility. The BDI-II was used for eligibility screening only and was not repeated as a baseline measure.

#### Personalized cue generation and ratings

Participants were asked to reflect on drug-related memories and verbally generate ten personalized cues capable of eliciting craving. They were instructed to generate specific cues rather than abstract or implicit cues, with examples provided to facilitate appropriate cue generation. Each cue was subsequently displayed on a computer screen and rated for arousal (excitement), salience (importance), valence (enjoyment), and vividness on a 5-point Likert scale (1 = not at all to 5 = very much), consistent with previous cue-rating procedures (29,35,49). For each participant, the six cues with the highest composite ratings across the four domains were selected for use during the intervention. Cues with a composite score below 12, equivalent to a mean rating below 3 across the four dimensions, were replaced by newly generated cues. The six selected personalized cues were then randomly assigned to the six prespecified temporal intervals within the participant’s allocated condition.

### Craving Measures

#### Phasic Craving

Phasic craving was assessed using the 13-item Desire for Drug Questionnaire (DDQ) (50), which measures current opioid craving across three subscales: (1) Desire and Intention to Drug Use, (2) Negative Reinforcement, and (3) Drug Abuse Control. Items are rated on a 7-point Likert scale (1 = strongly disagree to 7 = strongly agree), and subscale scores were expressed as mean item scores. Higher scores on the Desire and Intention to Drug Use and Negative Reinforcement subscales indicate greater craving-related responding, whereas higher scores on the Drug Abuse Control subscale indicate greater perceived control over drug use. The validated Persian version of the DDQ (51) was administered immediately before and after each of the three intervention sessions. The three DDQ subscales constituted the primary phasic-craving outcomes.

### Tonic Craving

Tonic craving was assessed using the 12-item Obsessive–Compulsive Drug Use Scale (OCDUS) (50), which assesses opioid craving over a broader time frame than the DDQ. The scale comprises four subscales: (1) Desire and Mental Preoccupation with Drugs, (2) Drug-Related Life Interference, (3) Motivation, Emotion, and Lack of Control, and (4) Resistance to Drug Use. Items are rated using five graded response options, and subscale scores were expressed as mean item scores on the 1–5 response scale. Higher scores generally indicate greater craving-related severity; for the Resistance to Drug Use subscale, however, higher scores indicate less effort to resist drug use. The validated Persian version of the OCDUS (51) was administered before the first intervention session and after the final intervention session to assess pre-to-post changes in tonic craving. The four OCDUS subscales constituted the primary tonic-craving outcomes.

### Delay Discounting Measure

Delay discounting was assessed using the 27-item Monetary Choice Questionnaire (MCQ) (52), which presents a series of choices between smaller-sooner and larger-later monetary rewards. The items comprise three sets of nine choices corresponding to small, medium, and large reward magnitudes. Responses were scored using the 27-Item Monetary Choice Questionnaire Automated Scorers (53), to estimate discounting parameters (*k*). Higher *k* values indicate steeper delay discounting and a stronger preference for immediate rewards. The geometric-mean *k* value was log-transformed for analysis; more negative log(*k*) values therefore indicate smaller *k* values and less steep delay discounting. A psychometrically validated Persian version of the MCQ (54) was administered before the first intervention session and after the final intervention session to assess pre-to-post changes in delay discounting. MCQ log(*k*) was the secondary outcome.

### Procedures

All study sessions were conducted individually by a trained cognitive psychologist in private clinical rooms. The first study session comprised eligibility screening, baseline assessment, personalized cue generation, and cue rating. This was followed by three intervention sessions, each lasting 75 minutes. Before each intervention session, urine screening was conducted for recent non-prescribed substance use.

At each intervention session, the DDQ was administered immediately before the intervention procedure began. Each session comprised two 30-minute intervention blocks separated by a 15-minute rest period. The first block began with a 5–8-minute stabilization period, followed by the active condition-specific intervention procedures. In the second block, the active intervention procedures were completed first, after which the DDQ was administered immediately and before the final 5–8-minute stabilization period. During each active intervention component, one personalized drug-related cue was presented in large, bold Persian text on a computer screen and used to initiate the condition-specific procedure. Participants’ emotional state and craving-related responses were monitored throughout cue exposure and the intervention exercises. Following the post-session DDQ, the final stabilization period was used to ensure that participants were calm and clinically stable before leaving the clinic. Participants were given an opportunity to report any distress or concerns and remained under researcher observation until they felt stable to leave.

The intervention structure was identical across the three sessions, with the principal difference between conditions being the temporal orientation of the episodic exercises. In CIREF, participants engaged with progressively more distant future time points (1 day, 1 week, 1 month, 3 months, 6 months, and 1 year), whereas participants in ERT engaged with recent-past time points (1, 2, 3, 5, 7, and 9 days before the session) (Figure 2). Two personalized cues, corresponding to two prespecified temporal intervals, were used during each intervention session. Thus, DDQ assessments were conducted immediately before the intervention and immediately after completion of the second active intervention component, before the final stabilization period. The OCDUS and MCQ were administered before the first intervention session and again after the final intervention session to assess pre-to-post changes in tonic craving and delay discounting, respectively.

**Figure 2.**
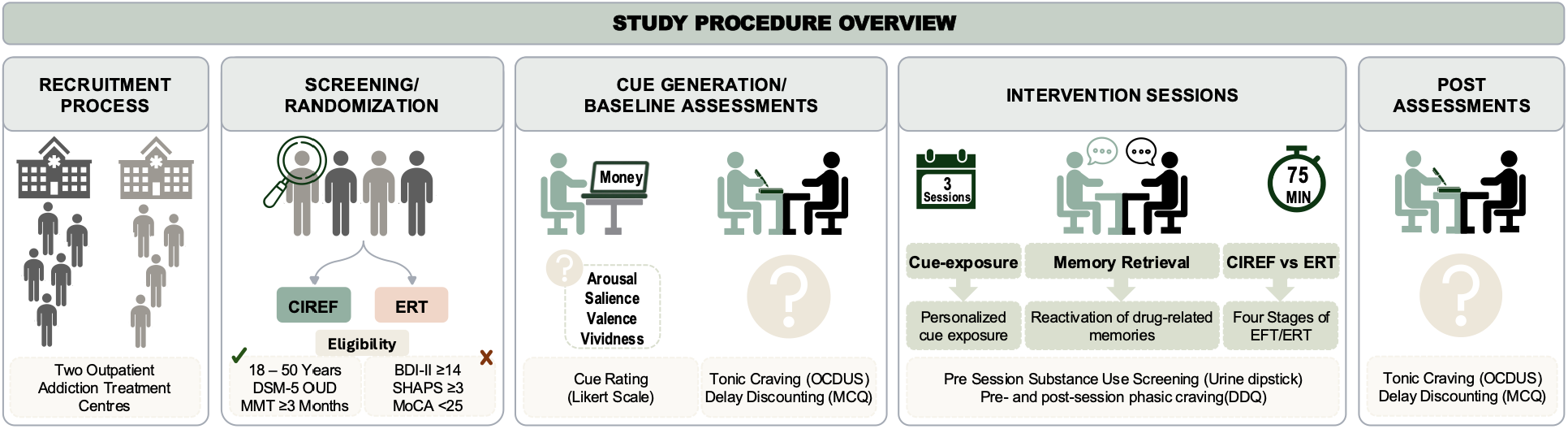
Study procedure overview of the CIREF pilot randomized controlled trial. Participants were recruited from two outpatient addiction treatment centres, screened for eligibility, and randomized to Cue-Induced Retrieval and Reconsolidation with Episodic Future Thinking (CIREF) or Episodic Recent Thinking (ERT). Personalized drug-related cues were generated and rated for arousal, salience, valence/enjoyment, and vividness, and six cues were selected for the intervention. Tonic craving (OCDUS) and delay discounting (MCQ) were assessed before the first intervention session and after the final intervention session. Participants completed three 75-minute intervention sessions, each including pre-session urine substance-use screening, personalized cue exposure and memory retrieval, condition-specific episodic exercises, and pre- and post-session assessment of phasic/current craving using the DDQ. **CIREF** = Cue-Induced Retrieval and Reconsolidation with Episodic Future Thinking; **ERT** = Episodic Recent Thinking; **BDI-II** = Beck Depression Inventory-II; **MoCA** = Montreal Cognitive Assessment; **SHAPS** = Snaith–Hamilton Pleasure Scale; **DDQ** = Desire for Drug Questionnaire; **OCDUS** = Obsessive–Compulsive Drug Use Scale; **MCQ** = Monetary Choice Questionnaire.

**Figure 3.**
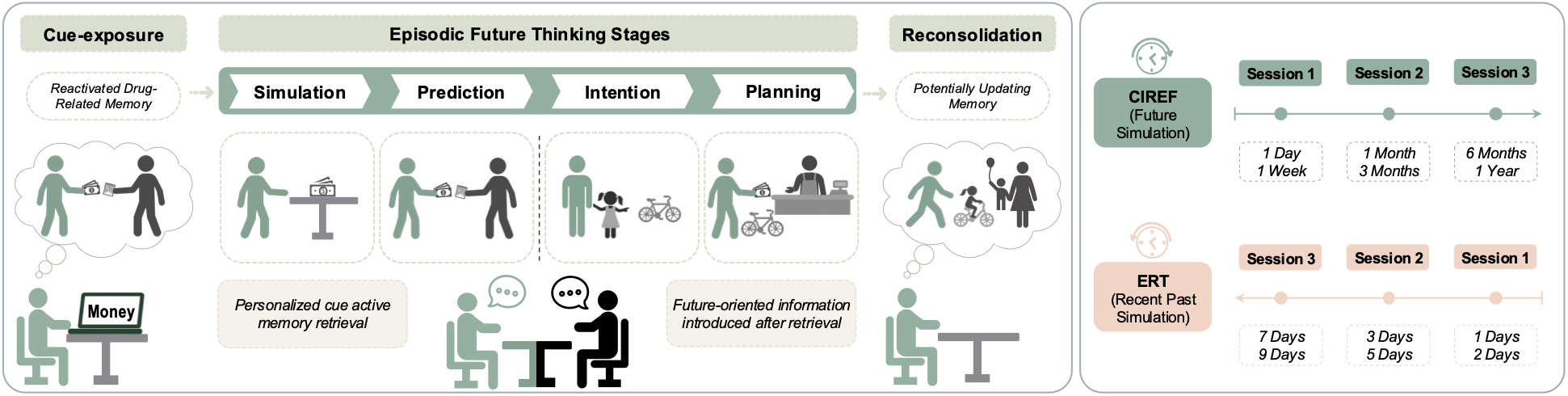
Conceptual structure and temporal organization of the CIREF and ERT interventions. Each intervention block began with exposure to a personalized drug-related cue and active retrieval of the associated drug-related memory. In CIREF, retrieval was followed by structured future-oriented processing comprising simulation, prediction, intention, and planning. Two future temporal intervals were used in each session: 1 day and 1 week in Session 1, 1 month and 3 months in Session 2, and 6 months and 1 year in Session 3. ERT followed the corresponding intervention structure but anchored episodic processing to the recent past, using intervals of 1 and 2 days in Session 1, 3 and 5 days in Session 2, and 7 and 9 days in Session 3. Within the CIREF framework, future-oriented information introduced after retrieval is hypothesized to provide content for potential retrieval-dependent memory updating; the present study did not independently establish memory destabilization or reconsolidation. **CIREF** = Cue-Induced Retrieval and Reconsolidation with Episodic Future Thinking; **ERT** = Episodic Recent Thinking.

### CIREF Intervention

Each CIREF intervention block began with cue-induced retrieval of a personalized drug-related memory. A selected personalized drug-related cue was presented as written text on a computer screen for approximately 1–2 minutes. Participants were instructed to view the cue and actively recall the associated drug-related memories. This retrieval procedure was intended to reactivate the corresponding drug-related memory representation and provide an opportunity to introduce new, personally relevant information under conditions potentially compatible with retrieval-dependent memory updating. Following cue-induced retrieval, the intervention proceeded through four stages derived from the taxonomy of future-oriented cognition proposed by Szpunar et al. (36):

#### *i)* Simulation

Simulation refers to the construction of a mental representation of a specific autobiographical future event (36). Participants were instructed to vividly simulate a hypothetical future event at the prespecified temporal interval in response to the presented drug-related cue. They were asked to project themselves into the imagined situation and verbally describe relevant episodic details including temporal, spatial, interpersonal, behavioural, and emotional context, for approximately 3 minutes. When necessary, the researcher used standardized prompts, such as “Where are you?”, “Who are you with?”, and “What are you doing or thinking?” to facilitate episodic elaboration.

#### *ii)* Prediction

Prediction refers to estimation of the likelihood of and/or one’s reaction to a specific autobiographical future event (36). Participants were instructed to anticipate possible reactions, emotions behavioural responses, and outcomes that might arise in the simulated drug-related situation within a 3-minute period. They were encouraged to imagine various strategies for managing potential drug-related situations in the future. While verbally expressing these predictions, participants were encouraged to consider both positive and negative scenarios that could result from exposure to drug- related cues. These anticipated responses and outcomes were then appraised in relation to their likely consequences.

#### *iii)* Intention

Intention refers to setting a goal in relation to a specific autobiographical future event (36). Following the prediction and appraisal, participants were asked to choose a specific, practical, and goal-directed intention for responding to the anticipated situation. The exercise encouraged consideration of more adaptive, self-controlled responses rather than choices dominated by immediate reward. This stage of intention formation resembles the “goal-setting exercises” commonly used in psychotherapy settings and the implementation of intentions (13,37), where participants clearly outline the details (when, where, and how) that will lead to goal attainment.

#### *iv)* Planning

Planning refers to organizing the steps required to arrive at a specific autobiographical future event (36). Participants were guided to plan and verbally articulate three concrete steps toward accomplishing their identified goal. Where appropriate, they were encouraged to include practical behavioural actions, including activities that involved physical engagement. (i.e., light running with a member of the family or friend). This stage was intended to translate future-oriented intentions into concrete actions and to introduce adaptive, goal-relevant information following retrieval of the drug-related memory.

Together, these four stages were designed to introduce personally relevant, future-oriented information following retrieval of drug-related memories and to strengthen prospective, goal-directed responses to drug-related cues. Within the CIREF framework, this future-oriented material is hypothesized to provide adaptive updating content that may become associated with reactivated drug-related representations (13). The present study did not independently establish memory destabilization or reconsolidation; accordingly, reconsolidation was treated as a proposed retrieval-dependent mechanism rather than an assumed consequence of the intervention.

### Active Control Intervention

Participants assigned to ERT received the same intervention dose, session structure, personalized cue-exposure procedures, therapist contact, and overall sequence of guided exercises as participants assigned to CIREF. The principal distinction was the temporal orientation of episodic processing. Rather than constructing future-oriented situations, ERT participants anchored the corresponding exercises to personalized cue-related experiences from the recent past, using intervals of 1, 2, 3, 5, 7, and 9 days before the session.

Following cue-induced retrieval, participants reconstructed the relevant recent-past cue-related event and described its temporal, spatial, interpersonal, behavioural, and emotional details. They then reflected on their responses, emotions, and outcomes associated with the recalled event, including alternative responses or outcomes where relevant, and completed corresponding intention and planning exercises anchored to that recent-past experience.

Thus, CIREF and ERT were matched on personalized cue exposure, memory retrieval, episodic elaboration, therapist contact, intervention duration, intention formation, planning, and general engagement with autobiographical information, while differing principally in temporal orientation: future-oriented episodic processing in CIREF versus recent-past episodic processing in ERT (55). Because the CIREF and ERT temporal schedules were not matched on absolute temporal distance, the design did not independently distinguish the effects of temporal direction from those of temporal horizon.

### Data Analysis

Primary analyses were conducted on the 28 participants who completed the intervention and post-intervention assessment. Statistical analyses were performed in Python 3.12 using pandas, NumPy, SciPy, Statsmodels, and Pingouin. Continuous variables are reported as M (SD) and categorical variables as n (%). Baseline continuous characteristics were compared using independent-samples t-tests (Student’s or Welch’s, as appropriate) or Mann–Whitney U tests when distributional assumptions were not met. Categorical variables were compared using chi-square tests or a Monte Carlo approximation to the Fisher–Freeman–Halton exact test for sparse contingency tables. Baseline comparisons were used descriptively to characterize the randomized groups rather than to evaluate the success of randomization. The primary craving outcomes comprised the mean scores of the three DDQ subscales assessing phasic craving and the mean scores of the four OCDUS subscales assessing tonic craving. MCQ log(*k*) was the secondary outcome.

For phasic craving, each of the three DDQ subscale mean scores was analyzed using a 2 (Group: CIREF, ERT) × 6 (Occasion) mixed-design ANOVA, with Occasion representing the six Session × Time assessments across the three intervention sessions. Holm correction was applied across the three DDQ Group × Occasion interaction tests. Mauchly’s test was used to assess sphericity, with Greenhouse–Geisser corrections applied when required. To characterize the temporal pattern of change, follow-up analyses decomposed Occasion into Group × Time (Pre vs. Post, averaged across sessions) and Group × Session (Sessions 1–3, averaged across Pre/Post assessments) effects.

For tonic craving, separate 2 (Group) × 2 (Time: Pre, Post) mixed-design ANOVAs were conducted for the mean scores of each of the four OCDUS subscales. Holm correction was applied across the four Group × Time interaction tests. Mann–Whitney U tests comparing pre-to-post change scores between groups were conducted as sensitivity analyses, with Holm correction applied across the four OCDUS subscales.

Delay discounting was analyzed using a 2 (Group) × 2 (Time: Pre, Post) mixed-design ANOVA on log-transformed MCQ discounting parameters [log(k)]. Follow-up analyses examined within-group pre-to-post changes and the between-group difference in change scores.

Effect sizes are reported as partial eta-squared (η²p) for ANOVA effects, Cohen’s d for independent-group comparisons, Cohen’s dz for paired comparisons, and rank-biserial correlation (r) for Mann–Whitney U tests, with 95% confidence intervals where applicable. All statistical tests were two-sided, with statistical significance set at p < .05. The implemented analyses differed from the OSF preregistration (GQTP7), including changes in model specification and multiplicity adjustment and the log transformation of MCQ k. These deviations are documented transparently in the protocol-amendment and study- history materials.

## RESULTS

### Baseline characteristics

Thirty participants were randomized to CIREF (n = 15) or ERT (n = 15). One participant in each group discontinued before completing the post-intervention assessment, resulting in 28 participants included in the analysis (CIREF, n = 14; ERT, n = 14; Figure 1). Baseline demographic, clinical, cognitive, and personalized cue-rating characteristics are presented in **Table 1**. No statistically significant between-group differences were observed for the assessed baseline characteristics (all ps ≥ .121). Ratings of the personalized drug-related cues (arousal, salience, valence, and vividness) were likewise comparable between groups. Consistent with recommendations for randomized pilot trials, these comparisons are presented descriptively to characterize the sample rather than as evidence of successful randomization (56).

**Table 1.** Baseline demographic, clinical, cognitive, and personalized cue-rating characteristics in CIREF and ERT groups.

| Variable | CIREF (n = 14) | ERT (n = 14) | Test statistic | p | Effect size |
| --- | --- | --- | --- | --- | --- |
| **Age (years)** | 33.86 (9.44) | 36.00 (7.58) | $t(26) = -0.66$ | .513 | $d = -0.25$ |
| **Occupation** | | | $\chi^2(1) = 0.00$ | 1.000<sup>a</sup> | |
| Employed | 12 (85.7) | 12 (85.7) |  |  |  |
| Unemployed | 2 (14.3) | 2 (14.3) |  |  |  |
| **Primary opioid used** | | | $\chi^2(2) = 4.27$ | .121<sup>a</sup> | |
| Natural opiates (e.g., opium) | 6 (42.9) | 10 (71.4) |  |  |  |
| Semi-synthetic opioids (e.g., heroin) | 0 (0.0) | 1 (7.1) |  |  |  |
| Multiple opioid use | 8 (57.1) | 3 (21.4) |  |  |  |
| **Route of administration (before treatment)** | | | $\chi^2(3) = 4.92$ | .179<sup>a</sup> | |
| Inhalation | 3 (21.4) | 6 (42.9) |  |  |  |
| Oral | 0 (0.0) | 2 (14.3) |  |  |  |
| Inhalation and oral | 9 (64.3) | 4 (28.6) |  |  |  |
| Intravenous | 2 (14.3) | 2 (14.3) |  |  |  |
| **Age at first opioid use (years)** | 17.71 (2.76) | 19.43 (4.36) | $t(26) = -1.24$ | .225 | $d = -0.47$ |
| **History of opioid use (years)** | 9.00 (6.39) | 9.64 (7.15) | $t(26) = -0.25$ | .804 | $d = -0.09$ |
| **Methadone dose (mg/day)** | 97.14 (28.60) | 91.79 (26.43) | $U = 110.5$ | .573 | $r = 0.13$ |
| **MoCA** | 27.00 (1.30) | 26.86 (1.17) | $U = 102.0$ | .862 | $r = 0.04$ |
| **SHAPS** | 1.50 (0.65) | 1.71 (0.61) | $U = 78.5$ | .329 | $r = -0.20$ |
| **Cue rating: Arousal** | 4.45 (0.70) | 4.38 (0.83) | $U = 92.5$ | .814 | $r = -0.06$ |
| **Cue rating: Salience** | 4.23 (0.74) | 4.39 (0.78) | $U = 77.5$ | .348 | $r = -0.21$ |
| **Cue rating: Valence (enjoyment)** | 4.29 (0.62) | 4.29 (0.76) | $U = 93.5$ | .852 | $r = -0.05$ |
| **Cue rating: Vividness** | 4.56 (0.44) | 4.52 (0.86) | $U = 85.0$ | .544 | $r = -0.13$ |

**Note.** Continuous variables are presented as M (SD) and categorical variables as n (%). Cohen's d is reported for independent-samples t tests and rank-biserial correlation (r) for Mann–Whitney U tests. Signed effect sizes are oriented as CIREF relative to ERT; positive values indicate higher values/ranks in CIREF and negative values indicate lower values/ranks in CIREF. MoCA and SHAPS values reported here were obtained during the post-eligibility baseline reassessment and were not used to re-determine eligibility. **CIREF** = Cue-Induced Retrieval and Reconsolidation with Episodic Future Thinking; **ERT** = Episodic Recent Thinking; **MoCA** = Montreal Cognitive Assessment; **SHAPS** = Snaith–Hamilton Pleasure Scale. <sup>a</sup> Monte Carlo approximation was used for categorical comparisons with expected cell counts < 5.

### Phasic Craving

The 2 (Group) × 6 (Occasion) mixed ANOVAs showed significant Group × Occasion interactions after Holm correction across the three DDQ subscales for Desire and Intention to Drug Use, FGG(1.23, 32.09) = 24.37, pHolm < .001, η²p = .484, and Negative Reinforcement, FGG(1.35, 35.23) = 17.67, pHolm < .001, η²p = .405, indicating greater reductions across assessments in CIREF than ERT. The Group × Occasion interaction for Drug Abuse Control was not significant, FGG(1.41, 36.64) = 1.91, pHolm = .172, η²p = .068. Significant Occasion effects were observed for all three subscales (all ps < .001). Decomposition showed significant Group × Session interactions for Desire and Intention, *F*GG(1.11, 28.97) = 26.20, *p* < .001, η²p = .502, and Negative Reinforcement, *F*GG(1.07, 27.73) = 21.34, *p* < .001, η²p = .451, but not Drug Abuse Control, *F*GG(1.20, 31.14) = 2.08, *p* = .157, η²p = .074. Group × Time (Pre/Post) interactions were likewise significant for Desire and Intention, *F*(1, 26) = 22.40, *p* < .001, η²p = .463, and Negative Reinforcement, *F*(1, 26) = 22.18, *p* < .001, η²p = .460, but not Drug Abuse Control, *F*(1, 26) = 0.37, *p* = .551, η²p = .014 (Table 2; Figure 4).

**Figure 4.**
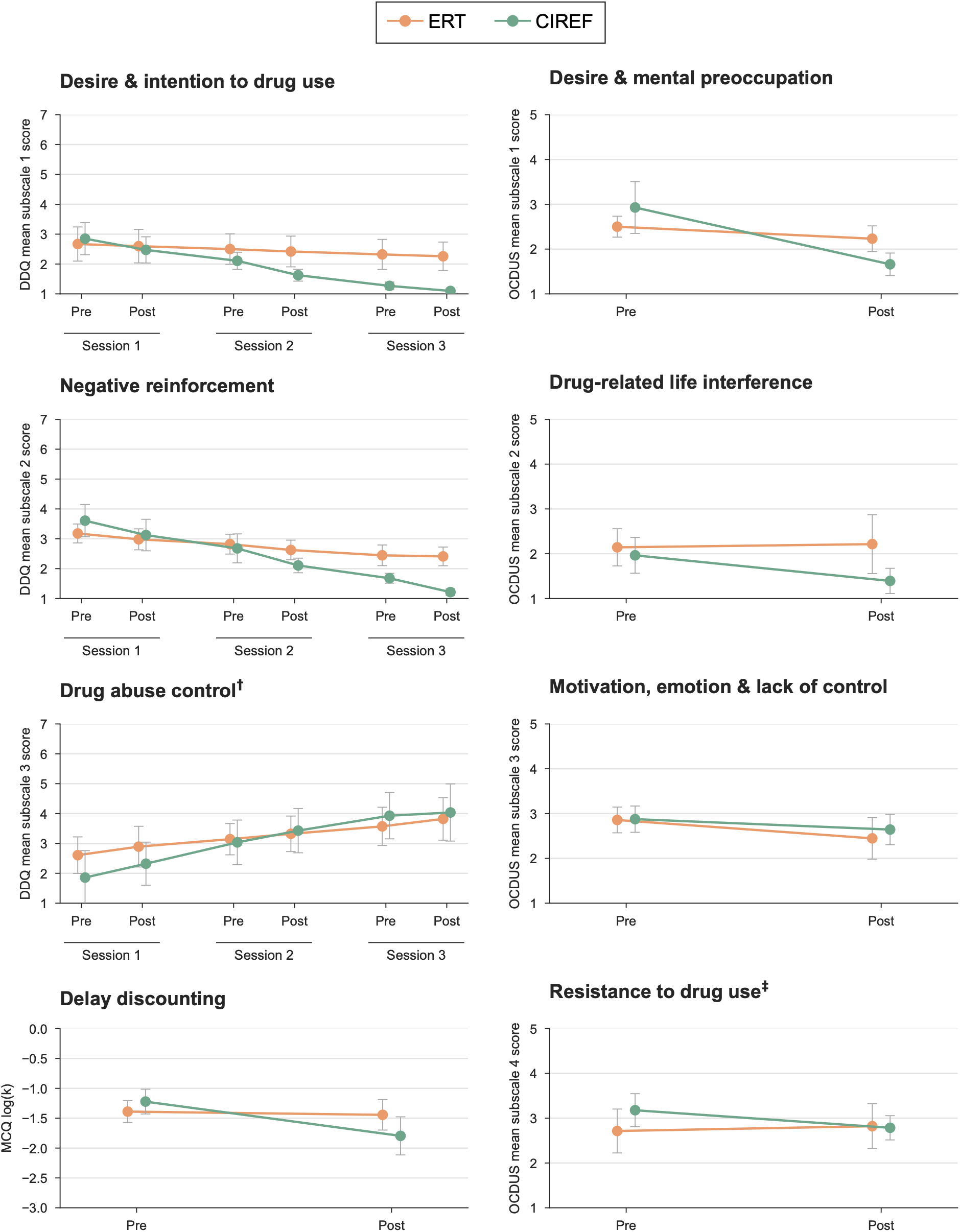
Changes in Phasic craving, tonic craving, and delay discounting in CIREF and ERT groups. Phasic craving was assessed using the three Desire for Drug Questionnaire (DDQ) subscales immediately before (Pre) and after (Post) each of the three intervention sessions. Tonic craving was assessed using the four Obsessive– Compulsive Drug Use Scale (OCDUS) subscales before the first and after the final intervention session. Delay discounting was assessed using Monetary Choice Questionnaire (MCQ) log(k) at the same pre- and post-intervention assessments. Data are shown for 28 participants (CIREF, n = 14; ERT, n = 14). Points represent observed group means; error bars indicate 95% confidence intervals. More negative MCQ log(k) values indicate smaller k and less steep delay discounting. **†** Higher DDQ Drug Abuse Control scores indicate greater perceived control over drug use. **‡** Higher OCDUS Resistance to Drug Use scores indicate less effort to resist drug use. **CIREF** = Cue-Induced Retrieval and Reconsolidation with Episodic Future Thinking; **ERT** = Episodic Recent Thinking; **DDQ** = Desire for Drug Questionnaire; **OCDUS** = Obsessive–Compulsive Drug Use Scale; **MCQ** = Monetary Choice Questionnaire.

**Table 2.** Desire for Drug Questionnaire subscale scores across intervention sessions and omnibus and decomposed mixed-design ANOVA results in CIREF and ERT groups

| DDQ subscale | Group | Session 1 | | Session 2 | | Session 3 | | Occasion | Group × Occasion | Group × Session | Group × Time |
| --- | --- | --- | --- | --- | --- | --- | --- | --- | --- | --- | --- |
| | | Pre | Post | Pre | Post | Pre | Post | FGG (df),<br>p, $\eta^2p$ | FGG (df),<br>$p_{Holm}$, $\eta^2p$ | FGG (df),<br>p, $\eta^2p$ | F<br>p, $\eta^2p$ |
| **Desire & Intention to Drug Use** | CIREF | 2.85<br>(0.93) | 2.47<br>(0.76) | 2.11<br>(0.50) | 1.62<br>(0.34) | 1.27<br>(0.22) | 1.10<br>(0.12) | 61.80(1.23,32.09),<br><.001, .704 | 24.37(1.23,32.09),<br><.001, .484 | 26.20(1.11,28.97),<br><.001, .502 | 22.40,<br><.001, .463 |
|  | ERT | 2.67<br>(0.99) | 2.60<br>(0.97) | 2.50<br>(0.89) | 2.42<br>(0.89) | 2.32<br>(0.87) | 2.26<br>(0.83) |  |  |  |  |
| **Negative Reinforcement** | CIREF | 3.61<br>(0.93) | 3.12<br>(0.91) | 2.68<br>(0.84) | 2.11<br>(0.42) | 1.68<br>(0.28) | 1.21<br>(0.19) | 70.91(1.35,35.23),<br><.001, .732 | 17.67(1.35,35.23),<br><.001, .405 | 21.34(1.07,27.73),<br><.001, .451 | 22.18,<br><.001, .460 |
|  | ERT | 3.18<br>(0.55) | 2.98<br>(0.61) | 2.82<br>(0.58) | 2.62<br>(0.57) | 2.45<br>(0.60) | 2.41<br>(0.54) |  |  |  |  |
| **Drug Abuse Control** | CIREF | 1.86<br>(1.56) | 2.32<br>(1.25) | 3.04<br>(1.29) | 3.43<br>(1.28) | 3.93<br>(1.34) | 4.04<br>(1.66) | 16.60(1.41,36.64),<br><.001, .390 | 1.91(1.41,36.64),<br>.172, .068 | 2.08(1.20,31.14),<br>.157, .074 | 0.37,<br>.551, .014 |
|  | ERT | 2.61<br>(1.06) | 2.89<br>(1.18) | 3.14<br>(0.91) | 3.32<br>(1.03) | 3.57<br>(1.11) | 3.82<br>(1.23) |  |  |  |  |

**Note.** Values are $M(SD)$ . DDQ subscale scores are mean item scores (range, 1–7). Occasion comprises the six sequential assessments (Session 1 Pre/Post, Session 2 Pre/Post, and Session 3 Pre/Post); Session represents the three intervention sessions averaged across Pre/Post assessments; Time represents Pre versus Post averaged across sessions. Greenhouse–Geisser (GG) corrected degrees of freedom are reported for Occasion and Session effects and their interactions with Group; Group × Time tests have $df = (1, 26)$ . For the primary Group × Occasion tests, $p_{Holm}$ denotes Holm-adjusted p values across the three DDQ subscales. The p values for the Group × Session and Group × Time decomposition analyses are unadjusted follow-up values. Higher Desire and Intention and Negative Reinforcement scores indicate greater craving, whereas higher Drug Abuse Control scores indicate greater perceived control over drug use. **CIREF** = Cue-Induced Retrieval and Reconsolidation with Episodic Future Thinking; **ERT** = Episodic Recent Thinking; **DDQ** = Desire for Drug Questionnaire; $\eta^2p$ = partial eta-squared.

### Tonic Craving

The 2 (Group) × 2 (Time) mixed ANOVAs showed a significant Group × Time interaction after Holm–Bonferroni correction across the four OCDUS subscales only for Desire and Mental Preoccupation with Drugs, F(1, 26) = 12.35, pHolm = .007, η²p = .322, 95% CI [.058, .534]. Mean scores decreased from 2.93 (1.00) to 1.66 (0.43) in CIREF, compared with 2.50 (0.40) to 2.23 (0.49) in ERT. Group × Time interactions for Drug-Related Life Interference, Motivation, Emotion, and Lack of Control, and Resistance to Drug Use were not significant after Holm–Bonferroni correction (adjusted ps ≥ .177). Mann–Whitney sensitivity analyses of pre-to-post change scores yielded the same pattern, with only Desire and Mental Preoccupation with Drugs remaining significant after Holm correction across the four subscales (pHolm = .005) (Table 3; Figure 4).

**Table 3.** pre- and post-intervention mean scores for the four Obsessive-Compulsive Drug Use Scale (OCDUS) subscale and Group × Time effects in CIREF and ERT groups

| OCDUS Subscales | Group | Pre<br>M(SD) | Post<br>M(SD) | Group × Time<br>F ($\eta^2_p$) | $p_{\text{Holm}}$ |
| --- | --- | --- | --- | --- | --- |
| Desire and Mental Preoccupation with Drugs | CIREF | 2.93 (1.00) | 1.66 (0.43) | 12.35 (.322) | .007 |
|  | ERT | 2.50 (0.40) | 2.23 (0.49) |  |  |
| Drug-Related Life Interference | CIREF | 1.96 (0.69) | 1.39 (0.49) | 3.69 (.124) | .177 |
|  | ERT | 2.14 (0.72) | 2.21 (1.14) |  |  |
| Motivation, Emotion, and Lack of Control | CIREF | 2.88 (0.51) | 2.64 (0.59) | 0.43 (.016) | .516 |
|  | ERT | 2.86 (0.50) | 2.45 (0.80) |  |  |
| Resistance to Drug Use | CIREF | 3.18 (0.64) | 2.79 (0.47) | 3.90 (.130) | .177 |
|  | ERT | 2.71 (0.85) | 2.82 (0.87) |  |  |

**Note.** Values are M (SD); $n = 14$ per group. OCDUS subscale scores are mean item scores on the 1–5 response scale. $p_{\text{Holm}}$ denotes Holm–Bonferroni-adjusted $p$ values across the four OCDUS subscale Group × Time interaction tests. Higher scores indicate greater craving-related severity, except for Resistance to Drug Use, for which higher scores indicate less effort to resist drug use. **CIREF** = Cue-Induced Retrieval and Reconsolidation with Episodic Future Thinking; **ERT** = Episodic Recent Thinking; **OCDUS** = Obsessive–Compulsive Drug Use Scale; $\eta^2_p$ = partial eta-squared.

### Delay Discounting

The 2 (Group) × 2 (Time) mixed ANOVA showed a significant Group × Time interaction for log-transformed MCQ *k* values, *F*(1, 26) = 7.18, *p* = .013, η²p = .217, 95% CI [.010, .446]. In the CIREF group, mean log(*k*) decreased from −1.22 (*SD* = 0.36) pre-intervention to −1.80 (*SD* = 0.55) post-intervention, indicating less steep delay discounting, whereas the ERT group showed little change (−1.39 [0.32] to −1.44 [0.44]). The pre-to-post effect was large in CIREF (*d*z = 0.99, 95% CI [0.34, 1.64]) and negligible in ERT (*d*z = 0.13, 95% CI [−0.40, 0.65]). The between-group difference in change scores was also significant, *t*(26) = 2.68, *p* = .013, *d* = 1.01, 95% CI [0.22, 1.80] (Table 4; Figure 4).

**Table 4.** Pre- and post-intervention MCQ log(k) scores and Group × Time interaction effect in CIREF and ERT groups.

| MCQ | Group | Pre<br>M(SD) | Post<br>M(SD) | Group × Time<br>F(df), $p$ $\eta^2_p$ |
| --- | --- | --- | --- | --- |
| MCQ $\log(k)$ | CIREF | -1.22 (0.36) | -1.80 (0.55) | 7.18(1, 26), .013, .217 |
|  | ERT | -1.39 (0.32) | -1.44 (0.44) |  |

**Note.** Values are M (SD); $n = 14$ per group. $\log(k)$ represents the log-transformed MCQ discounting parameter; more negative values indicate a smaller $k$ and less steep delay discounting. Because MCQ $\log(k)$ was the single secondary outcome, no multiplicity correction was applied to the Group × Time test. **CIREF** = Cue-Induced Retrieval and Reconsolidation with Episodic Future Thinking; **ERT** = Episodic Recent Thinking; **MCQ** = Monetary Choice Questionnaire; $\eta^2_p$ = partial eta-squared.

## DISCUSSION

This pilot randomized controlled trial provides the first empirical evaluation of Cue-Induced Retrieval and Reconsolidation with Episodic Future Thinking (CIREF) against a structurally matched Episodic Recent Thinking (ERT) active control in individuals with OUD receiving MMT. CIREF produced greater reductions in Desire and Intention to Drug Use and Negative Reinforcement, whereas Drug Abuse Control did not show a reliable differential effect after Holm correction across the three DDQ subscales. For tonic craving, only Desire and Mental Preoccupation with Drugs showed a reliable differential reduction after Holm correction across the four OCDUS subscales, whereas the other three dimensions did not. CIREF also produced a greater shift toward more negative MCQ log(*k*), corresponding to less steep monetary delay discounting. Overall, the craving effects were confined to dimensions involving desire/intention, negative-reinforcement motivation, and mental preoccupation rather than extending across all measured craving dimensions, alongside a shift toward less steep monetary delay discounting.

### Phasic Craving

The DDQ assesses momentary or current opioid craving, making the present findings most directly interpretable as changes in session-level current craving rather than enduring craving or cue reactivity per se (50,51). The greater reduction in Desire and Intention to Drug Use with CIREF suggests a reduction in the immediate motivational inclination to use opioids. This finding is compatible with a prospective-valuation account, whereby making delayed personal outcomes more accessible through episodic future simulation increases their influence on present evaluation and reduces the relative motivational weight of immediate rewards (25). However, the present study did not test whether changes in future-oriented valuation mediated changes in craving. Negative Reinforcement captures a distinct motivational component of craving—the tendency to use opioids to alleviate or escape unpleasant internal states—and its greater reduction could reflect reduced subjective value of opioid use as a relief response (50,57). This account remains inferential because EFT research has not established a specific effect on negative-reinforcement craving or this proposed relief-valuation pathway. Together, the two effects implicate related but distinguishable aspects of drug-use motivation: the immediate inclination to use and the anticipated relief associated with use (11,57). The absence of a reliable differential effect on Drug Abuse Control further indicates that the CIREF effect did not extend across all dimensions of current craving. Although this brief DDQ dimension has shown comparatively weaker psychometric performance in some substance-specific validation studies, this cannot be assumed to explain the present null finding (58,59).

Interpretation of the phasic-craving findings depends importantly on assessment timing: the DDQ was administered immediately before each intervention session and again after completion of both active intervention blocks, rather than after each individual personalized cue. The outcome therefore reflects session-level change in current craving rather than discrete cue-by-cue reactivity (60). The post-session DDQ was completed before the final stabilization period, so the subsequent stabilization procedure cannot account for the measured pre-to-post change in craving. CIREF and ERT received equivalent personalized cue-exposure doses and were broadly matched on intervention duration, therapist contact, episodic elaboration, and the general sequence of guided exercises; differences in cue-exposure quantity or nonspecific therapist contact are therefore less plausible as sole explanations for the differential effect. Repeated cue exposure can itself attenuate subjective craving through extinction or other exposure-related learning (61,62). but because this component was shared by both conditions, it cannot readily explain the greater reductions observed with CIREF.

One candidate explanation is that repeatedly pairing retrieval of drug-related representations with prospective representations of alternative consequences and actions altered cue-related expectancies or motivational significance in CIREF (13). However, neither expectancy nor cue-specific motivational significance was directly assessed, so this possibility cannot be distinguished from other perspective, affective, or learning-based processes. Because no independent measure of affective state was collected, transient condition-related emotional changes could also have influenced post-session self-reported craving (57,63). Accordingly, the findings support greater reductions in session-level self-reported Desire and Intention to Drug Use and Negative Reinforcement craving with CIREF relative to ERT, but do not provide direct evidence that CIREF reduced physiological cue reactivity or modified cue–drug associative responding itself.

### Tonic craving

The tonic-craving findings showed a similarly selective pattern, with a reliable differential reduction only in Desire and Mental Preoccupation with Drugs. This dimension captures drug-related desire and cognitive preoccupation over a broader time frame than the current craving assessed by the DDQ, whereas the remaining OCDUS dimensions encompass broader content related to interference with work and life, motivation, emotion and lack of control, and resistance to drug use (50,51). Drug-related cues can acquire heightened motivational salience and facilitate the accessibility of drug-related thoughts and motivational responses, providing one possible framework for understanding persistent desire and preoccupation with drug use (11,64,65). Within this framework, repeated cue retrieval followed by future-oriented processing in CIREF may have shifted the relative accessibility or motivational weighting of drug-related versus personally meaningful future representations; however, neither representational accessibility nor cue-specific motivational salience was directly assessed, so this remains a candidate explanation rather than an established mechanism (13,23,29). The convergent reductions in DDQ Desire and Intention to Drug Use and OCDUS Desire and Mental Preoccupation with Drugs suggest a cross-measure pattern involving desire-related content assessed over different temporal frames. The additional reduction in DDQ Negative Reinforcement extends this pattern to the immediate anticipated relief associated with opioid use (50,57). Together, the findings suggest a provisional cross-measure pattern centered on desire, preoccupation, and relief-related motivation rather than a generalized craving effect. Accordingly, the apparent convergence across craving dimensions should be regarded as hypothesis-generating and requiring replication, rather than evidence that CIREF selectively modifies an established underlying craving mechanism.

### Delay discounting

CIREF produced a greater shift toward more negative MCQ log(*k*) than ERT, corresponding to a smaller discounting parameter and less steep delay discounting.

Steeper delay discounting reflects greater devaluation of delayed outcomes and has been consistently associated with addictive behavior (66,67). This pattern is consistent with experimental evidence that episodic simulation of personally relevant future events can reduce delay discounting by increasing the influence of delayed outcomes on present intertemporal decisions (24,25,35). More broadly, a meta-analysis of 47 studies found a moderate overall effect of EFT on delay discounting (Hedges’ *g* = 0.52) (68), with EFT- related reductions also reported across several substance-using populations (27,29,30,69).

Convergent evidence also comes from a study of smokers in which EFT involving positive future events reduced delay discounting relative to ERT involving real events from the recent past (55). This supports consistency with the broader EFT-versus-ERT literature without establishing future temporal direction itself as the causal factor. In functional terms, CIREF may have broadened the temporal window over which consequences influenced present choice, increasing the relative weight of delayed outcomes in current intertemporal decisions (29). Importantly, this interpretation concerns the structured prospective processing implemented in CIREF—simulation, prediction, intention, and planning—rather than generic imagination alone (13,36). Because ERT received the same general intervention structure, including corresponding intention and planning exercises, unequal exposure to these components is less plausible as a sole explanation for the between-group MCQ difference. However, CIREF participants showed numerically steeper discounting at baseline, so chance baseline imbalance may have contributed to the magnitude of the observed pre-to-post difference in this small sample. Moreover, the MCQ assesses a domain conceptually proximal to the future-oriented processing trained in CIREF, making near transfer a plausible component of the observed effect. Accordingly, the present findings demonstrate change in hypothetical monetary intertemporal choice, not generalized impulsivity, opioid-specific valuation, or real-world delayed-reward behavior (52).

### CIREF as an integrated conceptual framework

Across outcomes, the observed pattern converged on aspects of drug-related desire, relief-related motivation, and preoccupation alongside the weighting of delayed relative to immediate outcomes, without extending across all measured craving dimensions. One candidate functional account is that CIREF may influence these outcomes through the coordinated engagement of several prospective processes rather than through a single unitary cognitive mechanism (13,36).

Together, simulation, prediction and appraisal, intention formation, and planning may make future consequences more accessible, support evaluation of alternative responses, and translate preferred responses into concrete prospective steps (13,24,25,36). These processes may contribute to craving and intertemporal choice through overlapping or distinct pathways; however, the present study did not measure mediation or experimentally isolate the individual intervention components. Accordingly, the data cannot establish a common temporal-window mechanism or a causal sequence in which changes in delay discounting produced changes in craving.

### Retrieval-Dependent Memory Updating

Each intervention block began with presentation of a personalized drug-related cue and active retrieval of the associated drug-related memory, followed by condition-specific episodic processing. However, the protocol did not include an experimentally designed prediction-error or expectancy-violation manipulation, nor an independent procedure intended to establish memory destabilization. This distinction is important because retrieval alone is not necessarily sufficient to destabilize a consolidated memory, and prediction error has been proposed as an important boundary condition influencing whether retrieval is followed by destabilization and subsequent reconsolidation (70,71). Accordingly, if cue-induced retrieval placed the drug-related memory into a modifiable state, the condition-specific episodic information introduced after retrieval could theoretically have contributed to retrieval-dependent updating (13,15,72). The present study included no measure or experimental manipulation that established memory destabilization, reconsolidation, modification of the original memory trace, or engagement of a reconsolidation window. The behavioural findings therefore cannot distinguish reconsolidation from other plausible processes, including prospective simulation, appraisal, new learning, inhibitory learning, expectancy change, or combinations of these processes. CIREF is consequently most appropriately characterized as a retrieval-based and reconsolidation-informed intervention, with reconsolidation remaining a proposed memory mechanism that requires direct experimental testing.

### ERT as an active control

The ERT condition represents an important design strength because the two conditions were matched on intervention duration, therapist contact, personalized cue presentation and memory retrieval, episodic elaboration, and the general sequence of guided exercises, including intention formation and planning. However, the episodic content itself differed: CIREF required construction of hypothetical future situations, whereas ERT required reconstruction of personalized cue-related experiences from the recent past. This matching makes the observed between-group differences less plausibly attributable to nonspecific episodic or autobiographical engagement, therapist attention, planning practice, or personalized cue exposure alone. However, the design does not isolate temporal direction from temporal distance, because CIREF extended from 1 day to 1 year into the future, whereas ERT remained within 1–9 days in the recent past. Consequently, future orientation and extended temporal horizon cannot be disentangled in the present comparison. Moreover, because future-oriented processing in CIREF was delivered immediately following personalized cue retrieval, the present comparison evaluates prospective processing within a retrieval-based intervention context rather than EFT administered in isolation. It therefore cannot determine whether the CIREF advantage arose from prospective processing itself or from its interaction with retrieval-dependent learning or updating. Accordingly, the defensible causal conclusion is that the CIREF intervention package as implemented produced greater changes than the matched recent-past ERT condition on selected outcomes—not that future temporal direction, EFT in isolation, temporal horizon, or reconsolidation independently caused those effects.

### CIREF as a Candidate Adjunct to Methadone Maintenance Treatment

MMT is an effective opioid-agonist pharmacotherapy for OUD, yet cue-related craving and steep delay discounting can remain clinically relevant among individuals receiving methadone maintenance treatment (73,74). By repeatedly engaging with personally salient drug-related situations and prospectively rehearsing anticipated consequences, preferred responses, intentions, and concrete action plans, CIREF may provide a complementary cognitive exercise alongside pharmacotherapy and support rehearsal of adaptive responses to situations that participants may subsequently encounter in everyday life (13,23,37). CIREF should therefore be considered a candidate adjunct designed to address proximal cognitive and motivational processes; whether these changes translate into clinically meaningful benefits, including reduced relapse or improved longer-term treatment outcomes, remains unknown.

### Limitations and future direction

Several limitations warrant consideration. First, the small male-only completer sample (n = 28) limits precision and may make effect-size estimates, particularly for individual craving dimensions, unstable. Larger, adequately powered trials with more diverse clinical samples are therefore needed. Second, analyses were restricted to completers and were not full intention-to-treat estimates of the randomized sample; future trials should prioritize all-randomized analyses, with per-protocol and missing-data sensitivity analyses as complementary approaches.

Third, outcomes were assessed only immediately after the intervention and were limited to self-reported craving and hypothetical monetary intertemporal choice. The study therefore does not establish durability or effects on physiological cue reactivity, opioid-specific valuation, opioid use, relapse, MMT retention or adherence, or broader functioning. Longer follow-up incorporating these clinical and behavioural outcomes is required. Fourth, the intervention provider was not blinded, fidelity was monitored but not quantitatively scored, and participants necessarily knew the temporal orientation of their exercises. Thus, procedural differences and expectancy or demand effects on self-report cannot be fully excluded. Future trials should include standardized independent fidelity ratings and assessment of treatment expectancy where feasible. Fifth, exclusion of participants with clinically relevant depressive symptoms, anhedonia, cognitive impairment, and active non-opioid substance use disorders limits generalizability to more heterogeneous MMT populations.

Several design questions also require direct testing. Future studies should match future- and past-oriented conditions on absolute temporal distance and counterbalance temporal horizons across sessions to separate temporal direction and distance from cumulative intervention exposure. Dismantling designs should estimate the contributions of cue-induced retrieval, episodic future processing, and their combination. To test reconsolidation specifically, experiments should manipulate proposed boundary conditions such as prediction error (20,71) and retrieval-to-intervention timing (15,72), while including independent measures of memory or cue reactivity rather than inferring destabilization from symptom change alone (21).

## CONCLUSION

In this pilot sample, CIREF produced greater changes than ERT in two dimensions of session-level phasic craving—Desire and Intention to Drug Use and Negative Reinforcement—one dimension of tonic craving, Desire and Mental Preoccupation with Drugs, and monetary delay discounting. These findings are preliminary and hypothesis-generating and support further investigation of CIREF as a retrieval-based, future-oriented intervention for individuals with OUD receiving MMT. They do not, however, establish memory reconsolidation, a single temporal-window mechanism, or effects on relapse, treatment retention, or sustained recovery.

## Data Availability

De-identified participant-level data and supporting materials may be made available upon reasonable request after publication of the primary study results, subject to review for scientific appropriateness, ethical acceptability, participant confidentiality, and feasibility. Approved requesters may be required to enter into a data-use agreement. Direct identifiers and potentially identifying free-text information, including personalized drug-cue content, will not be shared. Study documentation, non-identifying study materials, analysis code, and publication-related supporting materials are publicly available through the Open Science Framework repository at https://osf.io/q6w9d/.

https://osf.io/gqtp7/

## Ethic statement

All participants provided written informed consent prior to participation. All participants were fully informed about the study procedures, and their participation was voluntary. They were assured that their decision to participate or withdraw at any time would not affect the clinical care they were receiving. Participants were compensated with a gift card for their time and involvement. The study was approved by the Ethics Committees of the University of Tehran and the Institute for Cognitive Science Studies (ICSS) (Approval Code: IR.UT.IRICSS.REC.1400.011) and was conducted in accordance with the principles of the World Medical Association Declaration of Helsinki.

## Author contributions

MT: Conceptualization, Methodology, Investigation, Data curation, Formal analysis, Writing original draft.

KG: Investigation, Data curation, Writing – review & editing.

PR: Conceptualization, Methodology, Writing – review & editing. MS: Resources, Project administration, Investigation.

JV: Supervision, review & editing. HE: Supervision, review & editing.

TR: Supervision, Conceptualization, Methodology, Writing – review & editing, Visualization. All authors reviewed and approved the final manuscript.

## Acknowledgements

We gratefully acknowledge Professor Warren K. Bickel for his valuable conceptual input that contributed to the development of the CIREF framework. We thank all participants for their time and engagement in this study and the clinical staff at the participating centres for their assistance with recruitment and coordination.

## Primary Funding

This study received no funding.

## Clinical trial registration details

The trial was prospectively registered in the Iranian Registry of Clinical Trials (IRCT; IRCT20210804052079N1) and preregistered through OSF Registries (OSF Preregistration; DOI: 10.17605/OSF.IO/GQTP7).

