## Supplementary Materials for "Cue-Induced Retrieval and Reconsolidation with Episodic Future Thinking for Craving and Delay Discounting in Opioid Use Disorder: A Pilot Randomized Controlled Trial"

This supplementary material provides expanded statistical results for the primary craving outcomes and secondary delay-discounting outcome reported in the main article. **Table S1** presents the full omnibus and decomposed mixed-design ANOVA results for the three DDQ subscales; **Table S2** presents the full mixed-design ANOVA results for the four OCDUS subscales; and **Table S3** presents the full mixed-design ANOVA results for MCQ log(k).

**Table S1** Descriptive statistics and full omnibus and decomposed mixed-design ANOVA results for DDQ subscale scores across the three intervention sessions in the CIREF and ERT groups

| DDQ subscale | Group | Session 1 | | Session 2 | | Session 3 | | Group $F$(df) $p$, $\eta^2p$ | Occasion $F_{GG}$ (df) $p$, $\eta^2p$ | Group × Occasion $F_{GG}$ (df) $p$, $\eta^2p$ | Time $F$(df) $p$, $\eta^2p$ | Group × Time $F$(df) $p$, $\eta^2p$ | Session $F_{GG}$ (df) $p$, $\eta^2p$ | Group × Session $F_{GG}$ (df) $p$, $\eta^2p$ |
|---|---|---|---|---|---|---|---|---|---|---|---|---|---|---|
| | | **Pre** | **Post** | **Pre** | **Post** | **Pre** | **Post** | | | | | | | |
| Desire & Intention to Drug Use | CIREF | 2.85 (0.93) | 2.47 (0.76) | 2.11 (0.50) | 1.62 (0.34) | 1.27 (0.22) | 1.10 (0.12) | 4.37(1,26), .047, .144 | 61.80(1.23,32.09), <.001, .704 | 24.37(1.23,32.09), <.001, .484 | 52.27(1.26), <.001, .668 | 22.40(1,26), <.001, .463 | 67.50(1.11,28.97), <.001, .722 | 26.20(1.11,28.97), <.001, .502 |
| | ERT | 2.67 (0.99) | 2.60 (0.97) | 2.50 (0.89) | 2.42 (0.89) | 2.32 (0.87) | 2.26 (0.83) | | | | | | | |
| Negative Reinforcement | CIREF | 3.61 (0.93) | 3.12 (0.91) | 2.68 (0.84) | 2.11 (0.42) | 1.68 (0.28) | 1.21 (0.19) | 3.02(1,26), .094, .104 | 70.91(1.35,35.23), <.001, .732 | 17.67(1.35,35.23), <.001, .405 | 70.82(1,26), <.001, .731 | 22.18(1,26), <.001, .460 | 87.80(1.07,27.73), <.001, .772 | 21.34(1.07,27.73), <.001, .451 |
| | ERT | 3.18 (0.55) | 2.98 (0.61) | 2.82 (0.58) | 2.62 (0.57) | 2.45 (0.60) | 2.41 (0.54) | | | | | | | |
| Drug Abuse Control | CIREF | 1.86 (1.56) | 2.32 (1.25) | 3.04 (1.29) | 3.43 (1.28) | 3.93 (1.34) | 4.04 (1.66) | 0.11(1,26), .741, .004 | 16.60(1.41,36.64), <.001, .390 | 1.91(1.41,36.64), .172, .068 | 16.49(1,26), <.001, .388 | 0.37(1,26), .551, .014 | 17.89(1.20,31.14), <.001, .408 | 2.08(1.20,31.14), .157, .074 |
| | ERT | 2.61 (1.06) | 2.89 (1.18) | 3.14 (0.91) | 3.32 (1.03) | 3.57 (1.11) | 3.82 (1.23) | | | | | | | |

***Note.*** Values are presented as mean item scores, M (SD); n = 14 per group. DDQ subscale scores range from 1 to 7. For each DDQ subscale, the omnibus analysis used a 2 (Group: CIREF, ERT) × 6 (Occasion: Session 1 Pre, Session 1 Post, Session 2 Pre, Session 2 Post, Session 3 Pre, Session 3 Post) mixed-design ANOVA. Follow-up analyses decomposed Occasion into Group × Time (Pre vs Post, averaged across sessions) and Group × Session (Sessions 1–3, averaged across Pre/Post assessments) effects. Greenhouse–Geisser-adjusted degrees of freedom are reported for Occasion, Group × Occasion, Session, and Group × Session where applicable. For the primary Group × Occasion tests, $p_{Holm}$ denotes Holm-adjusted p values across the three DDQ subscales; p values for the Group × Time and Group × Session decomposition analyses are unadjusted follow-up values. Higher scores indicate greater craving-related severity for Desire and Intention to Drug Use and Negative Reinforcement, whereas higher Drug Abuse Control scores indicate greater perceived control over drug use. **CIREF** = Cue-Induced Retrieval and Reconsolidation with Episodic Future Thinking; **ERT** = Episodic Recent Thinking; **DDQ** = Desire for Drug Questionnaire; **$\eta^2p$** = partial eta-squared.

**Table S2** Pre- and post-intervention OCDUS mean subscale scores and mixed-design ANOVA results in the CIREF and ERT groups.

| OCDUS Subscales | Group | Pre M(SD) | Post M(SD) | Group F(1, 26), p, η²p | Time F(1, 26), p, η²p | Group × Time F(1, 26), η²p | *p*Holm |
|---|---|---|---|---|---|---|---|
| Desire and Mental Preoccupation with Drugs | CIREF | 2.93 (1.00) | 1.66 (0.43) | 0.14, .713, .005 | 29.12, <.001, .528 | 12.35, .322 | .007 |
| | ERT | 2.50 (0.40) | 2.23 (0.49) | | | | |
| Drug-Related Life Interference | CIREF | 1.96 (0.69) | 1.39 (0.49) | 4.01, .056, .134 | 2.24, .147, .079 | 3.69, .124 | .177 |
| | ERT | 2.14 (0.72) | 2.21 (1.14) | | | | |
| Motivation, Emotion, and Lack of Control | CIREF | 2.88 (0.51) | 2.64 (0.59) | 0.33, .572, .012 | 5.62, .025, .178 | 0.43, .016 | .516 |
| | ERT | 2.86 (0.50) | 2.45 (0.80) | | | | |
| Resistance to Drug Use | CIREF | 3.18 (0.64) | 2.79 (0.47) | 0.78, .386, .029 | 1.27, .270, .047 | 3.90, .130 | .177 |
| | ERT | 2.71 (0.85) | 2.82 (0.87) | | | | |

***Note.*** Values are presented as mean item scores, M (SD); n = 14 per group. OCDUS subscale scores are expressed on the 1–5 response scale. Group, Time, and Group × Time effects are from separate 2 (Group: CIREF, ERT) × 2 (Time: Pre, Post) mixed-design ANOVAs for each OCDUS subscale, with df = (1, 26) for all effects. pHolm denotes Holm-adjusted p values across the four Group × Time interaction tests. Higher scores indicate greater craving-related severity, except for Resistance to Drug Use, for which higher scores indicate less effort to resist drug use. **CIREF** = Cue-Induced Retrieval and Reconsolidation with Episodic Future Thinking; **ERT** = Episodic Recent Thinking; **OCDUS =** Obsessive-Compulsive Drug Use Scale; **η²p** = partial eta-squared.

**Table S3** Pre- and post-intervention MCQ log(k) scores and mixed-design ANOVA results in the CIREF and ERT groups.

| MCQ | Group | **Pre** M(SD) | **Post** M(SD) | **Group** F(1,26), p ($\eta^2_p$) | **Time** F, p ($\eta^2_p$) | **Group × Time** F, p ($\eta^2_p$) |
|---|---|---|---|---|---|---|
| **MCQ log(k)** | CIREF | −1.22 (0.36) | −1.80 (0.55) | *F*(1,26) = 0.51, *p* = .481, η²p = .019 | *F*(1,26) = 10.52, *p* = .003, η²p = .288 | *F*(1, 26) = 7.18, *p* = .013, η²p = .217 |
| | ERT | −1.39 (0.32) | −1.44 (0.44) | | | |

***Note.*** Values are presented as M (SD); n = 14 per group. Effects are from a 2 (Group: CIREF, ERT) × 2 (Time: Pre, Post) mixed-design ANOVA. MCQ log(k) represents the log-transformed discounting parameter; more negative log(k) values indicate smaller k values and less steep delay discounting. Because MCQ log(k) was the single secondary outcome, no multiplicity correction was applied to the Group × Time interaction test. CIREF = Cue-Induced Retrieval and Reconsolidation with Episodic Future Thinking; ERT = Episodic Recent Thinking; MCQ = Monetary Choice Questionnaire; η²p = partial eta-squared. **CIREF** = Cue-Induced Retrieval and Reconsolidation with Episodic Future Thinking; **ERT** = Episodic Recent Thinking; **MCQ** = Monetary Choice Questionnaire; **η²p** = partial eta-squared.

**Table S4** OCDUS change-score sensitivity analyses using Mann-Whitney U tests.

| OCDUS subscale | CIREF change median [IQR] | ERT change median [IQR] | U | Unadjusted p | pHolm | Rank-biserial r |
|---|---|---|---|---|---|---|
| **Desire and Mental Preoccupation with Drugs** | -1.12 [-1.75, -0.56] | -0.25 [-0.25, 0.00] | 27.5 | .001 | .005 | -0.72 |
| **Drug-Related Life Interference** | -0.25 [-1.00, 0.00] | 0.00 [-0.88, 0.50] | 65.0 | .124 | .364 | -0.34 |
| **Motivation, Emotion, and Lack of Control** | -0.12 [-0.75, 0.25] | -0.62 [-0.94, 0.00] | 116.5 | .403 | .403 | 0.19 |
| **Resistance to Drug Use** | -0.50 [-1.00, 0.38] | 0.00 [0.00, 0.00] | 65.0 | .121 | .364 | -0.34 |

***Note.*** Change is Post - Pre; therefore, negative values indicate pre-to-post reduction in the subscale score. Tests compare change-score distributions between CIREF and ERT. pHolm is Holm-adjusted across the four OCDUS sensitivity tests. Rank-biserial r is oriented as CIREF relative to ERT; negative values indicate lower (more negative) change-score ranks in CIREF. For Resistance to Drug Use, the substantive interpretation differs because higher scores indicate less effort to resist drug use. **OCDUS =** Obsessive-Compulsive Drug Use Scale.
